# Empowering PET Imaging Reporting with Retrieval-Augmented Large Language Models and Reading Reports Database: A Pilot Single Center Study

**DOI:** 10.1101/2024.05.13.24307312

**Authors:** Hongyoon Choi, Dongjoo Lee, Yeon-koo Kang

## Abstract

**Introduction:** The potential of Large Language Models (LLMs) in enhancing a variety of natural language tasks in clinical fields includes medical imaging reporting. This pilot study examines the efficacy of a retrieval-augmented LLM system considering zero-shot learning capability of LLMs, integrated with a comprehensive PET reading reports database, in improving referring previous reports and decision-making.

**Methods:** We developed a custom LLM framework enhanced with retrieval capabilities, leveraging a database encompassing nine years of PET imaging reports from a single center. The system employs vector space embedding of the reports database to facilitate retrieval based on similarity metrics. Queries prompt the system to retrieve embedded vectors, generating context-based answers and identifying similar cases or differential diagnoses from the historical reports database.

**Results:** The system efficiently organized embedded vectors from PET reading reports, showing that imaging reports were accurately clustered within the embedded vector space according to the diagnosis or PET study type. Based on this system, a proof-of-concept chatbot was developed and showed the framework’s potential in referencing reports of previous similar cases and identifying exemplary cases for various purposes. Additionally, it demonstrated the capability to offer differential diagnoses, leveraging the vast database to enhance the completeness and precision of generated reports.

**Conclusions:** The integration of a retrieval-augmented LLM with a large database of PET imaging reports represents an advancement in medical reporting within nuclear medicine. By providing tailored, data-driven insights, the system not only improves the relevance of PET report generation but also supports enhanced decision-making and educational opportunities. This study underscores the potential of advanced AI tools in transforming medical imaging reporting practices.

## INTRODUCTION

The integration of Large Language Models (LLMs) into the clinical domain has heralded a new era in healthcare innovation, particularly in the realm of medical imaging reports (*1,2*). LLMs, with their sophisticated zero-shot learning capabilities, have shown promise in parsing, summarizing, and generating complex medical texts, thereby enhancing the efficiency and accuracy of clinical documentation and decision-making processes (*3*). Their application extends across various specialties, aiming to revolutionize how healthcare professionals interact with and leverage vast amounts of medical data for patient care.

Despite the growing interest and proven benefits of LLMs in many areas of medicine, their potential has not been fully explored in the realm of nuclear medicine imaging, particularly PET imaging reporting. Despite the potential of ChatGPT to revolutionize content creation by generating human-like text (*4*), specific applications leveraging LLMs in the nuclear medicine field, particularly for imaging reports, has not been explored. PET imaging, which is performed for a variety of purposes and conditions, produces complex data requiring thorough analysis and interpretation, playing a critical role in clinical decision-making (*5,6*). There is a need for advanced tools to aid in referencing past reports, sourcing cases for educational purposes, and conducting differential diagnoses, especially as the use of PET, which encompasses various radiotracers and diseases, becomes more widespread. This unmet need presents a significant opportunity for LLMs to improve the specificity and relevance of PET report generation. By leveraging prior reports and analogous case studies, LLMs can provide clinicians with valuable insight, aiding them in making informed decisions.

In this study, we introduce a pioneering approach to PET imaging reporting by developing and accessing a custom-built, retrieval-augmented LLM framework (*7*). This system leverages a comprehensive large database of PET reading reports. By embedding these reports into a vector space for efficient retrieval based on similarity metrics, our framework aims to significantly improve the process of referencing previous similar cases, enhancing the accuracy of differential diagnoses and facilitating educational opportunities. This proof-of-concept study seeks to demonstrate the feasibility and benefits of integrating advanced LLM capabilities with a vast repository of PET imaging data, aiming to set enhanced medical imaging reporting practices.

## MATERIALS AND METHODS

### Dataset

This study was conducted at a single center, utilizing reading reports of PET imaging data sourced from the clinical data warehouse (CDW) of the SUPREME Platform. We extracted data spanning from 2010 to 2023, comprising reports from 118,107 patients across 211,813 cases. Institutional Review Board (IRB) approval was secured from our hospital (IRB No. 2401-090-1501), with the requirement for written informed consent waived due to the retrospective nature of the study and the use of deidentified information. The dataset encompassed reading reports for all cases, along with the exam date, exam name, a deidentified research identifier (ID), sex, and date of birth (year-month format).

### Model architecture

In this study, we designed a proof-of-concept chatbot system for efficiently querying reading reports from a substantial dataset. It was based on ‘Retrieval-Augmented Generation (RAG)’ (*7*). The adaptability of this system allows for the utilization of various database formats, including but not limited to ‘csv’ files, to accommodate different sources of reading reports. This system amalgamates state-of-the-art language model technologies with sophisticated natural language processing and information retrieval techniques, aiming to deliver precise, contextually relevant responses to inquiries concerning PET imaging reading reports. The overall workflow of this system is illustrated in **Figure 1**.

**Figure 1:**
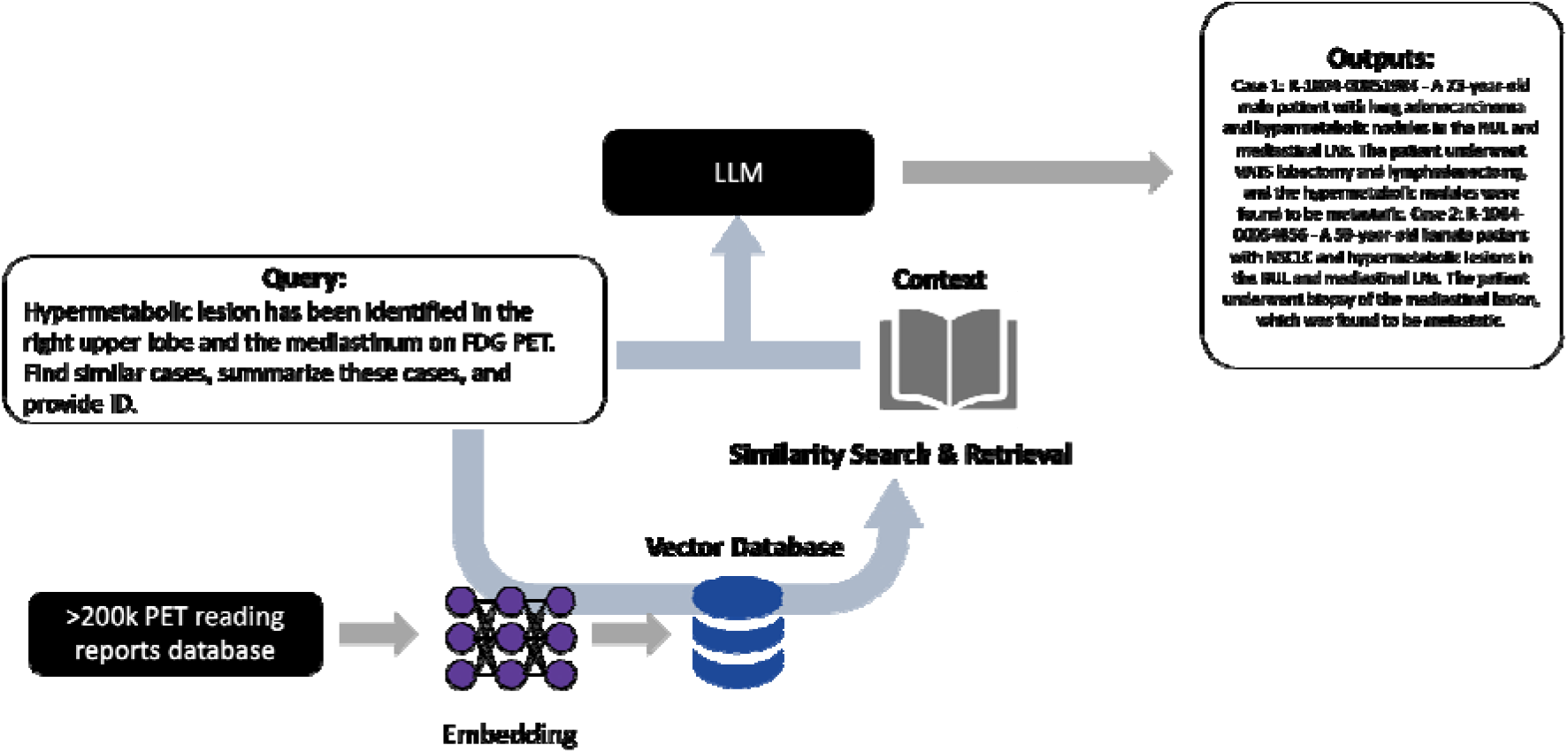
Workflow of the Chatbot System for Querying PET Imaging Reading Reports. The overall workflow of the proof-of-concept system designed for efficient querying of reading reports from a substantial dataset is illustrated. The system integrates the Retrieval-Augmented Generation (RAG) model with advanced language model technologies, natural language processing, and information retrieval techniques. The workflow demonstrates the process from user query input through to the delivery of the relevant reading report, showcasing the operational framework and interaction with different sources of reading reports.

The architecture of our system is underpinned by a series of modular components, each crucial for interpreting and responding to user queries. At the forefront is a sentence embedding layer, crafted to process intricate texts and queries by transforming sentences into vectors. This transformation facilitates subsequent processing by various mathematical models. We employed the Sentence Transformer model, specifically the “paraphrase-multilingual-MiniLM-L12-v2” (https://huggingface.co/sentence-transformers/paraphrase-multilingual-MiniLM-L12-v2), renowned for its ability to comprehend and paraphrase texts across multiple languages—a necessary feature considering the bilingual nature (English and Korean) of the reading reports in our dataset. To manage and retrieve PET reading reports effectively, our system incorporates a vector storage mechanism, Chroma (Chroma, https://www.trychroma.com/). Chroma organizes textual data into a searchable vector space by converting text into numerical vectors derived from the sentence embeddings. This conversion enables the system to execute advanced retrieval operations, identifying responses that are semantically relevant to the queries posed.

A pivotal element of our system is the integration of a question-answering (QA) component that synergizes the capabilities of the language model and vector storage system. This QA mechanism is adept at comprehending user queries, sourcing the most pertinent documents from the dataset, and formulating informative responses that precisely address the queries. For the generation of these responses, we incorporated the Llama-2 language model (*8*) and the system architecture was based on Langchain (*9*).

### Visualization of vector embedding

Following the process of sentence embedding, the resulting vectors were stored in a vector database. These vectors played a crucial role in identifying similarities between various texts, including the queries submitted to the system. To facilitate a deeper understanding of how PET reading reports are represented within this vector space, we employed t-distributed Stochastic Neighbor Embedding (t-SNE) for visualization purposes (*10*). Specific keywords associated with imaging reports, such as “lung cancer,” “breast cancer,” “lymphoma,” “methionine PET,” and “PSMA PET,” were chosen for this analysis. The objective was to ascertain whether reports containing these selected terms would naturally form distinct clusters within the vector space. This approach aimed to visually demonstrate the effectiveness of our vector embedding process in grouping similar reports, thereby providing insights into the semantic relationships and similarities between different PET reports in the dataset.

### Test examples

In the evaluation of prototype chatbots designed for navigating an extensive database of PET reading reports, we focused on testing their ability to accurately retrieve reports similar to those specified in user queries and to assist in differential diagnosis by referencing previous reports. This involved assessing the proficiency in identifying cases with specific diagnoses or imaging findings and their capability to extract relevant information to support nuclear medicine experts in diagnosing complex cases. The testing protocol simulated real-world scenarios, presenting the chatbots with diverse clinical questions to comprehensively evaluate their utility in clinical decision-making and their effectiveness in leveraging the vast database to enhance the accuracy and relevance of their responses.

## RESULTS

We analyzed PET imaging reports from 118,107 patients, totaling 211,813 cases, by converting them into vector embeddings. These embeddings were then visualized on a t-SNE plot to demonstrate dimensionality reduction and the clustering of reports with similar characteristics (**Figure 2A**). Each point on this plot represents a unique PET imaging report, with a specific case highlighted in red for illustrative purposes, including its original report. To evaluate the representational efficacy of the embeddings, reports containing key diagnostic terms such as ‘lung cancer’, ‘breast cancer’, and ‘lymphoma’, as well as those pertaining to specific types of exams like ‘C-11 methionine PET’ and ‘Ga-68 PSMA-11 PET’, were marked on the plot. The formation of distinct clusters on the t-SNE plot (**Figure 2B**) underscores the embeddings’ capability to effectively reflect the similarity among cases, affirming the method’s potential in facilitating the identification and visualization of related PET imaging reports.

**Figure 2.**
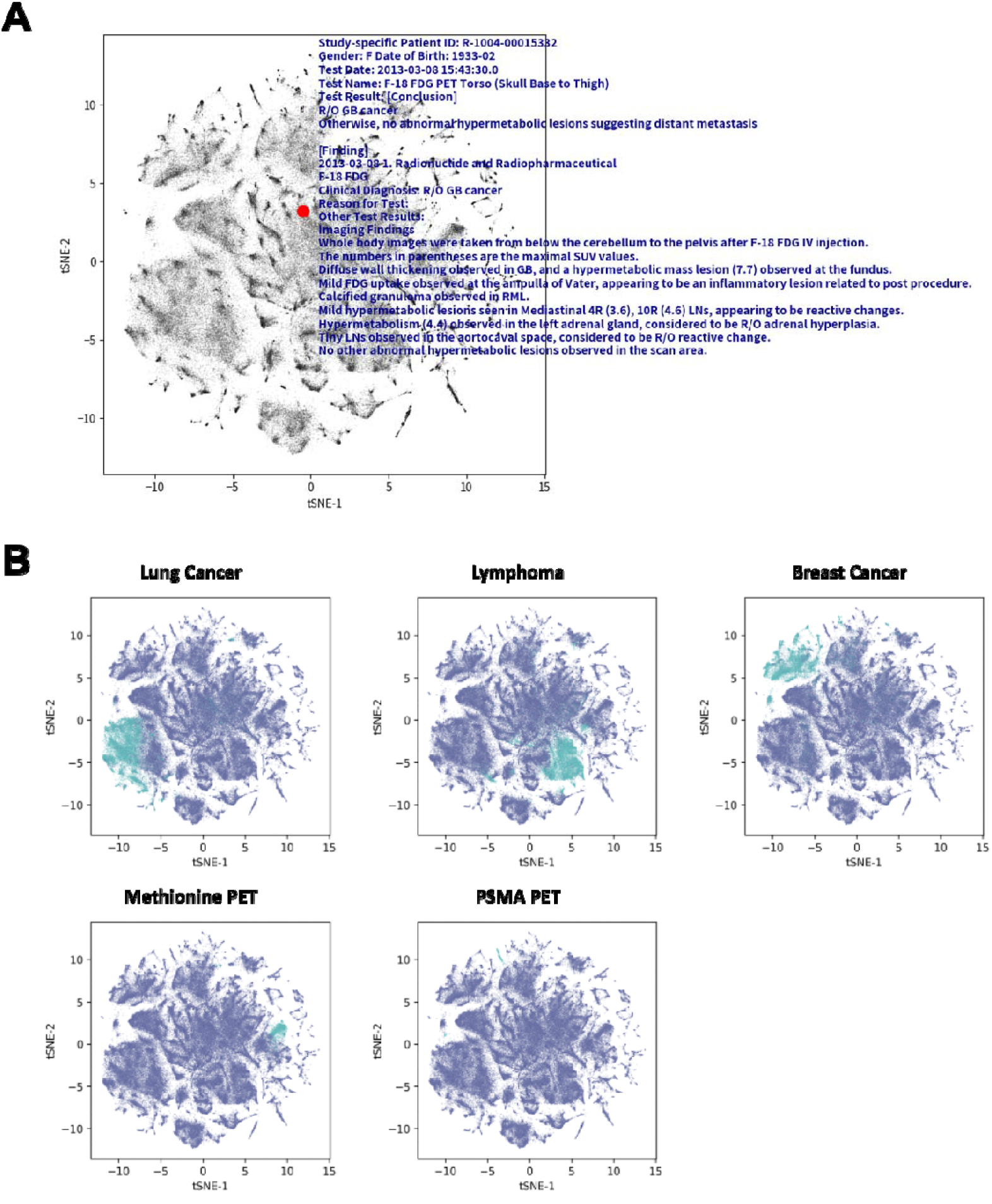
Visualization of PET Imaging Report Embeddings Using t-SNE. (A) t-SNE plot illustrates PET imaging report embeddings from 118,107 patients, totaling 211,813 cases. Each point on the plot represents a unique report, with a selected case highlighted in red to show an example of an original report. (B) t-SNE plots showcases the clustering efficacy of the embeddings, highlighting how reports containing key diagnostic terms like ‘lung cancer’, ‘breast cancer’, ‘lymphoma’, and specific types of exams such as ‘C-11 methionine PET’ and ‘Ga-68 PSMA-11 PET’ form distinct clusters. These clusters indicate the embeddings’ capability to reflect the similarity among cases, demonstrating the potential of this method in facilitating the identification and visualization of related PET imaging reports.

Using the prototype chatbot, we tested its efficacy in identifying cases pertinent to specific user queries. A notable instance involved the chatbot’s response to the query, “*Identify cases of breast cancer with metastasis to internal mammary lymph nodes*,” where it proficiently located and presented relevant cases from the database of prior reading reports (**Figure 3A**) (More examples are presented in **Supplementary Video 1**). Additionally, we evaluated the chatbot’s functionality in offering differential diagnoses by leveraging its integration with LLM. This was exemplified in a scenario where the chatbot was tasked to provide differential diagnoses for the condition described as “*Multiple mediastinal lymph nodes with increased FDG uptake without an identified primary site*.” The chatbot responded with a detailed list of differential diagnoses, accompanied by reference identifiers, enabling medical professionals to compare the suggested cases with their clinical outcomes and final diagnoses (**Figure 3B**).

**Figure 3:**
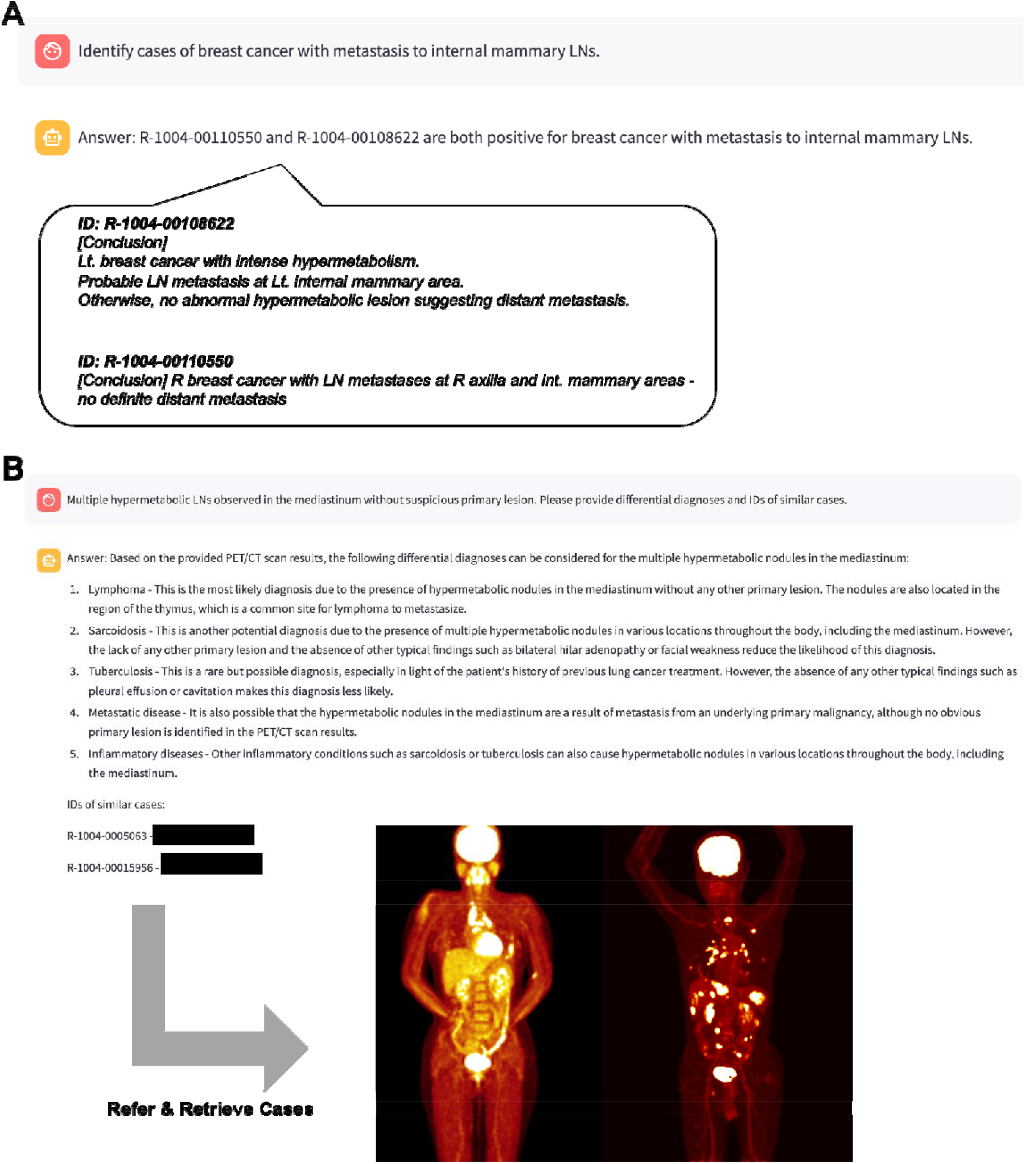
Examples of Chatbot Responses to Queries. (A) An example case displays an instance of the chatbot’s capability to accurately identify and present relevant cases in response to a user query about breast cancer with metastasis to internal mammary lymph nodes. It highlights the capacity to navigate a vast database of previous reading reports to identify relevant cases. (B) An example of the utility of system in generating differential diagnoses is displayed. This is demonstrated through the chatbot’s response to a query, where it offers a detailed list of potential diagnoses along with reference identifiers. As an example, by employing identifiers within the PACS system (in this example, we used deidentified information), prior imaging cases could be referenced for understanding cases and supporting decision making.

## DISCUSSION

In this study, we have explored the integration of LLMs into the PET imaging reporting process, presenting a novel prototype chatbot based on RAG capable of retrieving relevant cases and offering differential diagnoses based on specific user queries. This RAG-based language model represents a feasibility for medical purposes in nuclear medicine imaging field, particularly by incorporating contextual understanding from previous PET imaging reports to respond to queries from nuclear medicine physicians. This approach marks a departure from simple chatbot functionalities, introducing a system that integrates with the clinical workflow to provide contextually relevant information and insights. This proof-of-concept not only validates the utility of LLMs in enhancing the PET reporting process but also underscores the potential of AI-assisted tools to augment diagnostic accuracy and clinical decision-making in nuclear medicine.

The RAG model combines the strengths of information retrieval and generative AI to offer precise and informative answers to complex medical queries. It works by first retrieving relevant documents or data points from a vast database—in this case, a collection of PET imaging reports. Following this, the model uses the retrieved information as a context to generate responses that are not only relevant but also enriched with the specificity and detail required for decision-making. This method allows the system to provide answers that are deeply informed by historical cases and existing medical knowledge, thereby supporting physicians in diagnosing and managing patient care with a higher degree of accuracy and confidence. In contrast to earlier language models that concentrated on singular tasks (*11-13*), models based on the RAG framework with LLMs can handle diverse queries and produce varied outputs. The RAG model, distinct from LLMs that rely solely on their pre-trained datasets, actively incorporates pertinent historical information during its response generation. Primarily, employing LLMs like ChatGPT or Gemini directly is constrained by their inability to access individual center databases, which restricts their reference to prior cases and clinical outcomes. Moreover, due to stringent regulations concerning clinical data and privacy, the transfer of clinical records to external AI servers is considered highly sensitive and is inherently prohibited in numerous healthcare institutions (*14,15*). In this context, implementing a RAG-based LLM framework that utilizes PET reading reports could address these challenges by facilitating the application of real-world data in each hospital, while also avoiding the various data-related regulatory constraints. In addition, this feature is especially beneficial in specialty fields like nuclear medicine, where insights drawn from previous cases are helpful for informed decision-making in current clinical scenarios.

The application of our system extends beyond diagnostic support, serving as a valuable educational resource. By facilitating access to similar cases, it enables medical practitioners and trainees to explore diverse clinical scenarios, thereby enhancing their diagnostic skills and understanding of nuclear medicine (*4,16*). Furthermore, the ability of this system to reference previous cases when providing differential diagnoses enriches the educational content with practical, real-world examples, fostering critical thinking and decision-making skills among trainees.

A significant advantage of our approach is its ability to correlate imaging findings with follow-up clinical results, including final diagnoses and clinical outcomes. By referring historical data-driven context into the imaging interpretation process, it could provide an opportunity to refer a holistic view of clinical journey of similar case, from the imaging to the final outcome (*17,18*). This comprehensive approach facilitates a more nuanced understanding of the potential implications of specific imaging findings, guiding physicians in crafting PET imaging interpretation that are informed by both the current condition and comparable past cases. The insights derived from this analysis are invaluable for informing differential diagnosis, predicting patient outcomes, and even anticipating potential complications. Such insights are crucial for bridging the gap between imaging findings and patient management strategies, ultimately contributing to improved patient care.

However, the study also acknowledges certain limitations, including the inherent risk of generating inaccurate information (hallucinations) and the current model’s reliance on textual data (*19,20*). Addressing these challenges and exploring the integration of multimodal data, such as combining visual and textual analysis, are identified as essential steps forward. This future direction promises not only to mitigate the limitations but also to further enrich the system’s utility by providing a more holistic approach to medical query answering and decision support.

## CONCLUSION

In conclusion, our suggested AI framework affirm the transformative potential of AI-assisted tools in nuclear medicine, particularly in the context of PET imaging report analysis. The integration of a retrieval-augmented LLM with a comprehensive PET imaging report database marks an advancement in the field of nuclear medicine, particularly in imaging interpretation reporting. This approach enhances the workflow of nuclear medicine physician and relevance of PET report generation, facilitating superior decision-making and providing educational benefits. It underscores the role in improving the quality and efficacy of medical care within nuclear medicine. Furthermore, as we look to the future, the development of visual-language combined models stands as a pivotal next step in overcoming current limitations and fully realizing the benefits of AI in medical imaging. This proof-of-concept study and suggested framework showed the feasibility of innovation, promising to enhance the quality of nuclear medicine imaging reporting through improved diagnostics, education, and patient management.

## Data Availability

All data produced in the present study are available upon reasonable request to the authors

## Acknowledgment

We employed ChatGPT, developed by OpenAI, exclusively for grammatical corrections and enhancements in clarity. ChatGPT did not generate any new content.

## Author Contributions

Conceptualization and design: H.C..; data acquisition: H.C. and Y.K.; data analysis: H.C. and D.J.L.; original draft preparation: H.C. and Y.K..; review and editing: H.C.; supervision: H.C.; funding acquisition: H.C., and Y.K. All authors have read and agreed to the submission of the manuscript.

## Ethics approval

The retrospective analysis of human data and the waiver of informed consent were approved by the Institutional Review Board of the Seoul National University Hospital (No. 2401-090-1501).

